# Association between intimate partner violence and modern contraceptive use among women presenting for abortion care in Lusaka, the capital city of Zambia

**DOI:** 10.64898/2026.07.30.26359339

**Authors:** Eugine Kaunda, Alice Hazemba, Patrick Kaonga, J.Anitha Menon, Ketty Mwansa Lubeya, Namaipo Nankamba, Maureen Masumo, Bellington Vwalika

**Affiliations:** Department of Obstetrics & Gynaecology, School of Medicine, University of Zambia, Lusaka, Zambia; Department of Community & Family Medicine, School of Public Health, University of Zambia, Lusaka, Zambia; Department of Epidemiology & Biostatistics, School of Public Health, University of Zambia, Lusaka, Zambia; Department of Health Promotion and Education, School of Public Health, University of Zambia, Lusaka, Zambia; School of Psychology, Curtin University, Dubai, United Arab Emirates; Department of Midwifery & Women’s Health, School of Nursing Sciences, University of Zambia, Lusaka, Zambia

**Author notes:** Corresponding author (EK). These authors contributed equally to this work (Eugine Kaunda, Alice Hazemba, Patrick Kaonga, J.Anitha Menon, Bellington Vwalika). These authors also contibuted equally to this work (Ketty Mwansa Lubeya, Namaipo Nankamba, Maureen Masumo).

**Keywords:** Intimate partner violence, modern contraceptives, abortion

## Abstract

Intimate Partner Violence (IPV) has been shown to affect women’s access to sexual and reproductive health services, including modern contraceptives. However, there is conflicting data in the literature on how the experience of IPV among women affects modern contraceptive use, with some studies showing decreased rates of modern contraceptive use, while others have shown increased rates of modern contraceptive use among these women. This study therefore, aimed at studying the association between the experience of IPV and modern contraceptive use among women presenting for abortion care in Lusaka District, Zambia.

A cross-sectional study was conducted from 1 November to 31 December 2024 at four hospitals in Lusaka District. Women seeking abortion care were enrolled via convenience sampling, and data were obtained using a pre-tested interviewer-administered questionnaire. The outcome variable was modern contraceptive use in the preceding six months; exposures included IPV within the prior six months, as well as socio-demographic and reproductive characteristics. Analyses comprised descriptive statistics, chi-square test and multiple logistic regression.

A total of 372 women participated in the study and data was analysed for 367 women, with a median age of 25 years (IQR 22-30). IPV was reported by 231 participants (62.9%), most commonly emotional violence (44.7%). Modern contraceptive use within the preceding six months was reported by 150 women (40.9%). The odds of modern contraceptive use was lower amon women with IPV exposure compared with those who were not exposed (aOR 0.3; 95% CI 0.17–0.52), but higher among women seeking abortion care services from Chawama First Level Hospital compared to those seeking services from the Women and Newborn University Teaching Hospital ((aOR 4.4; 95% CI 1.68 – 11.65) and among women aged above 24 years compared to adolescent girls and young women (AGYW) aged 14-24 years (aOR 14; 95% CI, 6.40 – 30.60).

More than half of the women accessing abortion care services in Lusaka district experienced intimate partner violence, and less than half had used modern contraceptives within six months before they participated in the study. Thus, policies and programmes aimed at increasing modern contraceptive use among women should pay particular attention at addressing IPV and the reproductive health needs of AGYW and women residing in peri-urban areas.

## Introduction

The United Nations Sustainable Development Goal number 3 has among it’s targets the goal of achieving universal access to sexual and reproductive health-care services, including family planning by 2030 [1]. Expanding access to modern contraceptives has numerous health benefits including reduction in maternal and under five child mortality and reductions in unintended pregnancies and unsafe abortions [2]. Many factors including the experience of Intimate Partner Violence (IPV) affect modern contraceptive use among women [3, 4]. The forms of IPV which have been widely studied are physical, sexual and emotional violence.

However, economic or financial form of IPV is increasingly being recognized as another form of IPV which may have similar consequences as the other three forms of IPV [5]. In one study, economic IPV was referred to as the “hidden or invisible” form of IPV [6]. Globally it is estimated that 27% of women experience physical or sexual Intimate Partner Violence [7]. In Zambia, the prevalence of IPV experienced by women of reproductive age stands 47% [8]. Women who experience IPV may have limited access to modern contraceptives and may be at increased risk of unsafe abortion [9, 10].

Though most studies on modern contraceptive use among women who experience IPV have shown that the experience of IPV is associated with a reduction in modern contraceptive use [11–13], some studies have shown increased use of modern contraceptives among women who experience it [14, 15]. A study which was done by the World Health Organization in 15 Low and Middle-Income Countries (LMICs) found that women who experienced IPV were 1.7 times more likely to have unintended pregnancy and 2.7 times more likely to have an abortion compared to women who did not experience IPV [16]. In Bangladesh, a facility-based cross-sectional study which was conducted among women seeking abortion services found that women’s experience of IPV was associated with reduced modern contraceptive use [17]. According to the Zambia Demographic and Health Survey (ZDHS) of 2018, 48% of married women aged 15-49 years were using modern contraceptives, with 54% of the users using injectables followed by those who were using oral contraceptive pills (OCPs) and implants each at 17% and the remaining 12% were using male condoms, intrauterine devices and female sterilisation [8]. However, it has not been explored as to whether the use of these modern contraceptive methods is different among women seeking abortion care services in Lusaka district. Previous studies have shown that socio-demographic characteristics are associated with modern contraceptive use. In one study involving young women in Sub-Sahara Africa, adolescents aged 15-19 years were found to have higher odds of unmet need for modern contraception compared to young women aged 20-24 years [18]. In another study which was done in Benin, modern contraceptive use among adolescent girls and young women (AGYW) aged 15-24 years was found to be significantly lower compared to women aged 25 years and above [19]. Modern contraceptive use has been found to be higher among women with higher educational levels compared to those with lower educational levels, employed women compared with unemployed women and married women compared with unmarried women [20–22]. Religion has equally been found to be associated with modern contraceptive use. In Bangladesh for instance, modern contraceptive use was found to be lower among muslim women compared to non-muslim women [21].

There is limited data regarding modern contraceptive use among women who experience IPV in Zambia. This study therefore aimed at studying the proportions of diferrent forms of IPV and the association between experience of IPV and modern contraceptive use among women presenting for abortion care in Lusaka district of Zambia. Understanding this association will allow for development of policies and strategies aimed at increasing modern contraceptive use among women who experience IPV.

## Materials and methods

### Study settings

An Institution-based cross-sectional study was conducted from 1st November to 31st December 2024 among women seeking abortion care services at four government owned hospitals in Lusaka district, the capital city of Zambia. Lusaka district has an estimated population of 2.2 million people [23] and it comprises both urban and peri-urban areas with varying socioeconomic characteristics, providing a diverse sampling frame. Participants were recruited from the Women and Newborn University Teaching Hospital (WNUTH) and three First-Level Hospitals (FLHs)namely, Chawama FLH, Kanyama FLH and Chipata FLH. These hospitals were selected to represent diverse socioeconomic contexts which included low-income high density areas and medium to high-income low density areas.

### Definitions

Some definitions of variables and terms were adopted from existing sources, while others were generated or re-categorised as necessary for our study-specific objectives.

#### Modern contraceptive methods

Scientifically proven products or medical procedures that effectively prevent pregnancy by interfering with reproduction from acts of sexual intercourse, and they include oral contraceptives pills, condoms, injectables, implants, sterilization and intrauterine devices [24]

#### Modern contraceptive use

Use of a modern contraceptive method regularly within the last 6 months before the current aborted pregnancy at the time of conception. If a participant indicated that they had used more than one modern contraceptive method (e.g oral contraceptive pills and condoms), only the most effective method which was used on a regular basis was noted in the dataset.

#### Intimate Partner Violence

Refers to behaviour within an intimate relationship that causes physical, sexual, psychological or economic harm.

#### Economic abuse

A type of domestic violence that includes controlling someone’s access to money and other resources, withholding financial information such as family income, or preventing someone from going to work or getting further education [25].

### Sample size determination and sampling procedures

The Cochran formula was used to culculate the overall sample size using single proportion formula by considering the following assumptions: p – proportion of contraceptive use among women seeking abortion (p = 41.3%) [26]; margin of error (5%) and Z-score at 95% confidence score (1.96). Considering 10% non-response rate, the final sample size was 382.

The sample size for each of the four study sites was determined proportionally based on three months flow of patients seeking abortion services. Convenience sampling was used to enroll participants until the desired sample size for each site was reached.

### Data collection instruments and procedures

A pre-tested interviewer administered questionnaire was used to collect the data. The questionnaire was pilot tested with ten participants, and minor modifications were made to improve clarity and cultural appropriateness. The Abuse Assessment Screen (AAS) tool [27] was adapted and used to screen for physical, sexual and emotion IPV, while the Economic Abuse Screening Tool (EAST) [25] was adapted and used to screen for economic IPV. The two screening tools were merged into one and incorporated in the questionnaire (Appendix 1). The questionnaire was initially prepared in English and translated into Nyanja and Bemba then back to English to check consistency. A woman was considered to have experienced IPV if she answered “yes” to any of the screening questions from the adapted AAS and EAST tools. Interviews were carried out after participants had received abortion care services and were stable to participate in the study. Participants were also informed about the availability of services for Gender Based Violence (GBV) at all the four study sites and that they were free to visit them if they needed any services.

### Data analysis

The data were entered on an excel spreadsheet and thereafter exported into Stata version 15 for analysis. The dependent variable was modern contraceptive use within the last 6 months, while independent variables were the experience of IPV, Socio-demographic and reproductive characteristics. Descriptive statistics were used to describe the data. Chi square tests were used to test the association between categorical independent variables and the outcome variable.

Independent variables that were significantly associated with the outcome variable were included in the logistic regression model. Multiple logistic regression was used to further assess the relationship between modern contraceptive use and independent variables with results being presented as crude and adjusted odds ratios with corresponding 95% confidence intervals. A p-value less than 0.05 was considered significant. Multicollineality was checked to see the correlation among independent variables. Model fitness was checked with Hosmer Lemeshow test with a p-value of greater than 0.05 being considered a good fit.

### Missing data

The only variable in which some data were missing was partner’s age. Five out of 372 women (1.3%) had a missing value on the “partner’s age” variable. Listwise deletion (complete case analysis) was applied as data were missing completely at random and only 1.3% in the affected variable.

### Ethical considerations

The study protocol was approved by the Zambian National Health Research Authority (Reference: NHRA-1395/21/07/2024), the University of Zambia Biomedical Research Ethics Committee (Reference: 5491-2024), the Lusaka Provincial Health Office and the managements of the four hospitals where the study was conducted. Informed consent and consent plus assent for women aged below 18 years was obtained from all participants prior to their participation in the study. Furthermore, participants aged below 18 years, just like adults were interviewed in the absence of their parents/guardians to uphold confidentiality. Participants were not compensated for participation in the study, but they were given transport refunds for a bus fare.

## Results

### Socio-demographic characteristics

A total of 372 women attending abortion care services at the four study sites paticipated in this study with a response rate of 97%. We excluded 5 women who had missing data on partner’s age, leaving 367 for analysis. The median age of participants was 25 years (IQR 22-30), while that of their partners was 30 years (IQR 26-36). Less than half (n=156, 42.5%) were AGYW aged 14-24 years, with the rest being older women aged 25 years and above (n=211, 57.5%). More than half (n=205, 56%) had attained secondary level of education, while about a quarter (n=90, 24.5%) had no education or had attained primary education, with about 20% (n=72) having attained tertiary education. Less than two-thirds (n=220, 60%) were married, while slightly less than one-third (n=112, 30.5%) were employed. Two-thirds (n=243, 66.2%) resided in high density areas, while only 6% (n=22) resided in low density areas. Slightly more than half (n=197, 53.7%) were Pentecostal Christians, with less than one-third (n=110, 30%) being Protestants and only 15.7% (n=57) being Catholics. More than half (n=206, 56.1%) were multiparous with 43.1% (n=161) being nulliparous (Table 1).

**Table 1:**
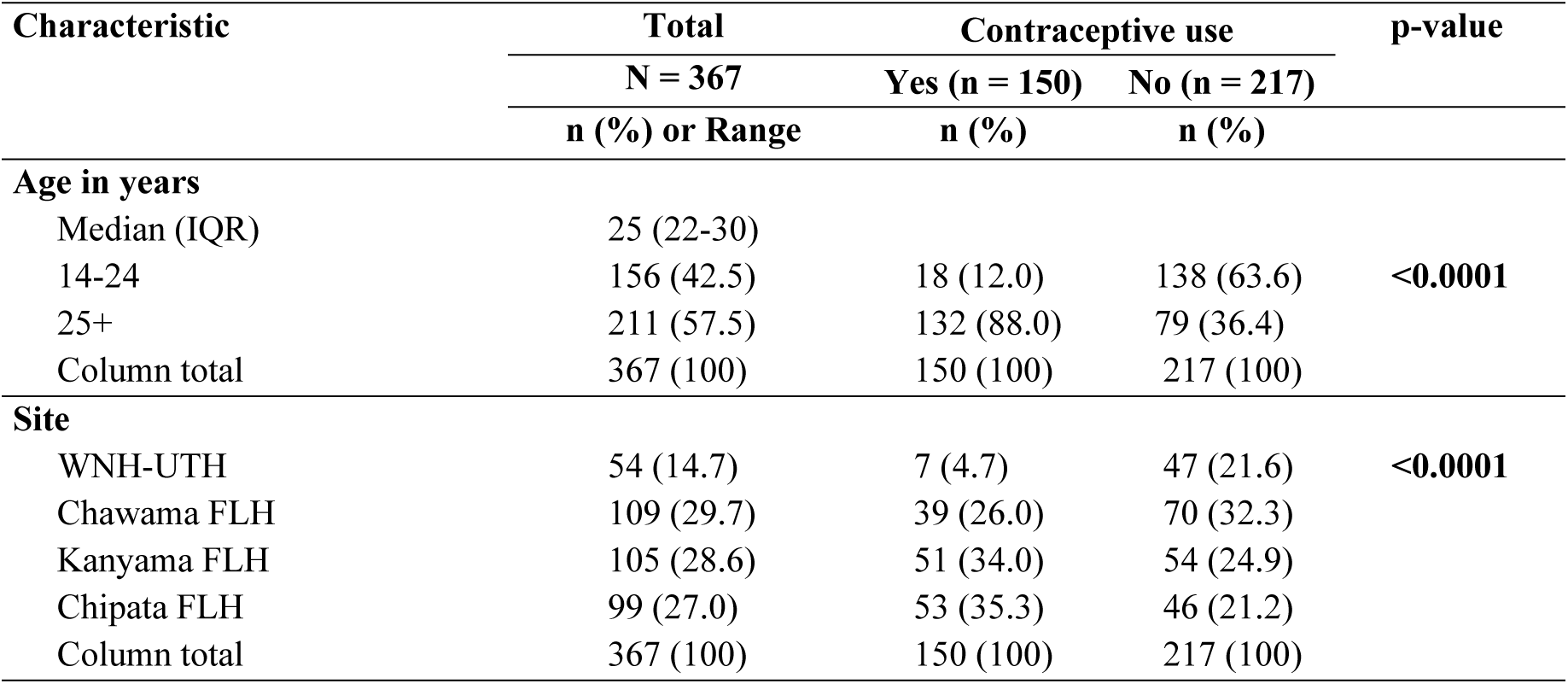
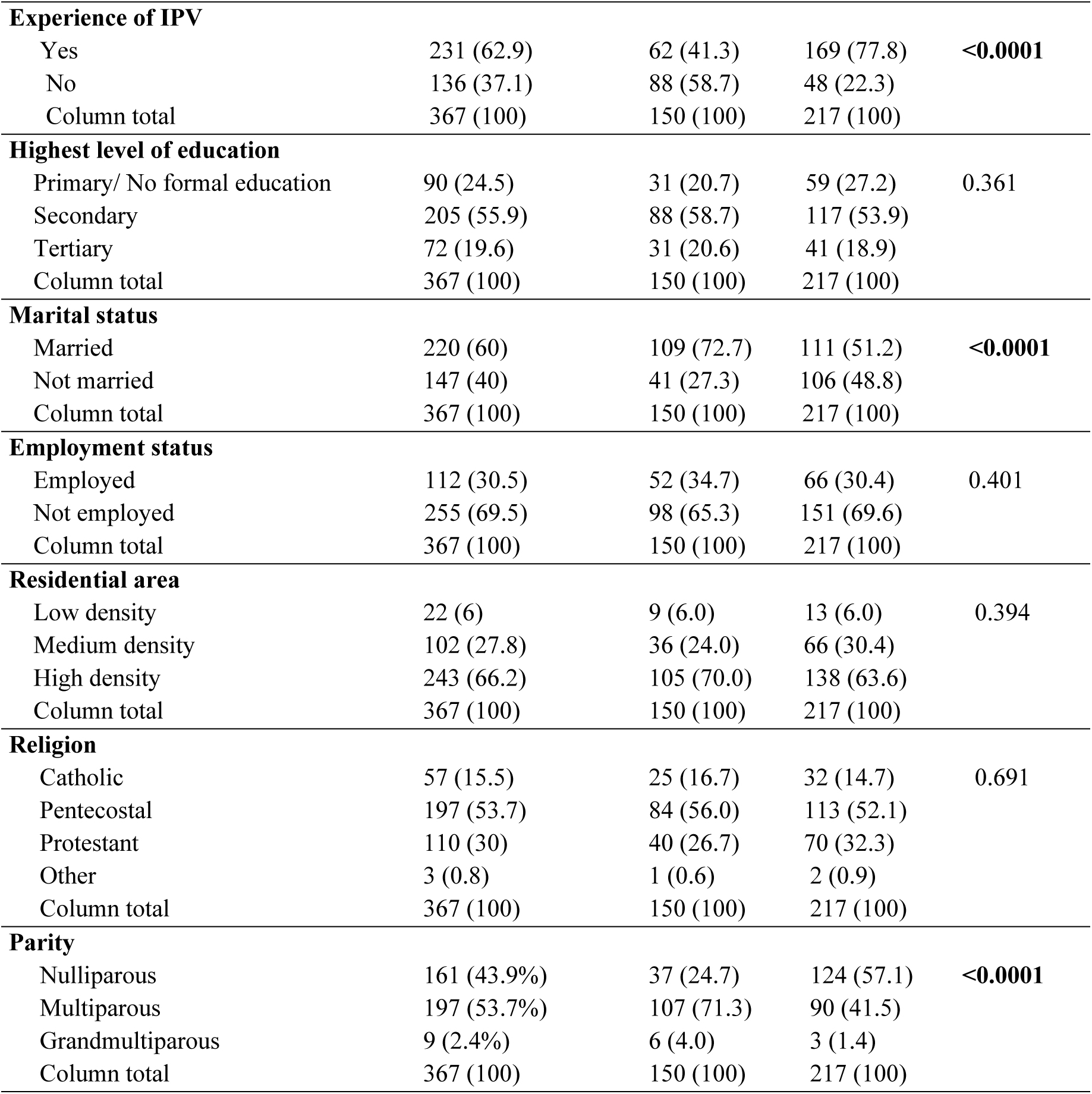
Socio-demographic, reproductive characteristics and bivariate analysis (Chi-square) of women seeking abortion care at the four selected hospitals in Lusaka district, Zambia, 2024 (n=367).

### Experience of Intimate Partner Violence

Slightly less than two-thirds (n=231, 62.9%) of the participants admitted having experienced one or more forms of IPV in the 6 months preceding the study. Of the 231 participants who experienced IPV, multiple responses regarding the forms of IPV experienced were possible resulting in a total of 412 responses to the forms of IPV experienced. Emotional violence was the commonest form of IPV experienced at 44.7% (n=184) followed by physical violence at 23.5% (n=97), economic violence at 20.1% (n=83) and the least was sexual violence at 11.7% (n=48).

**Fig 1.**
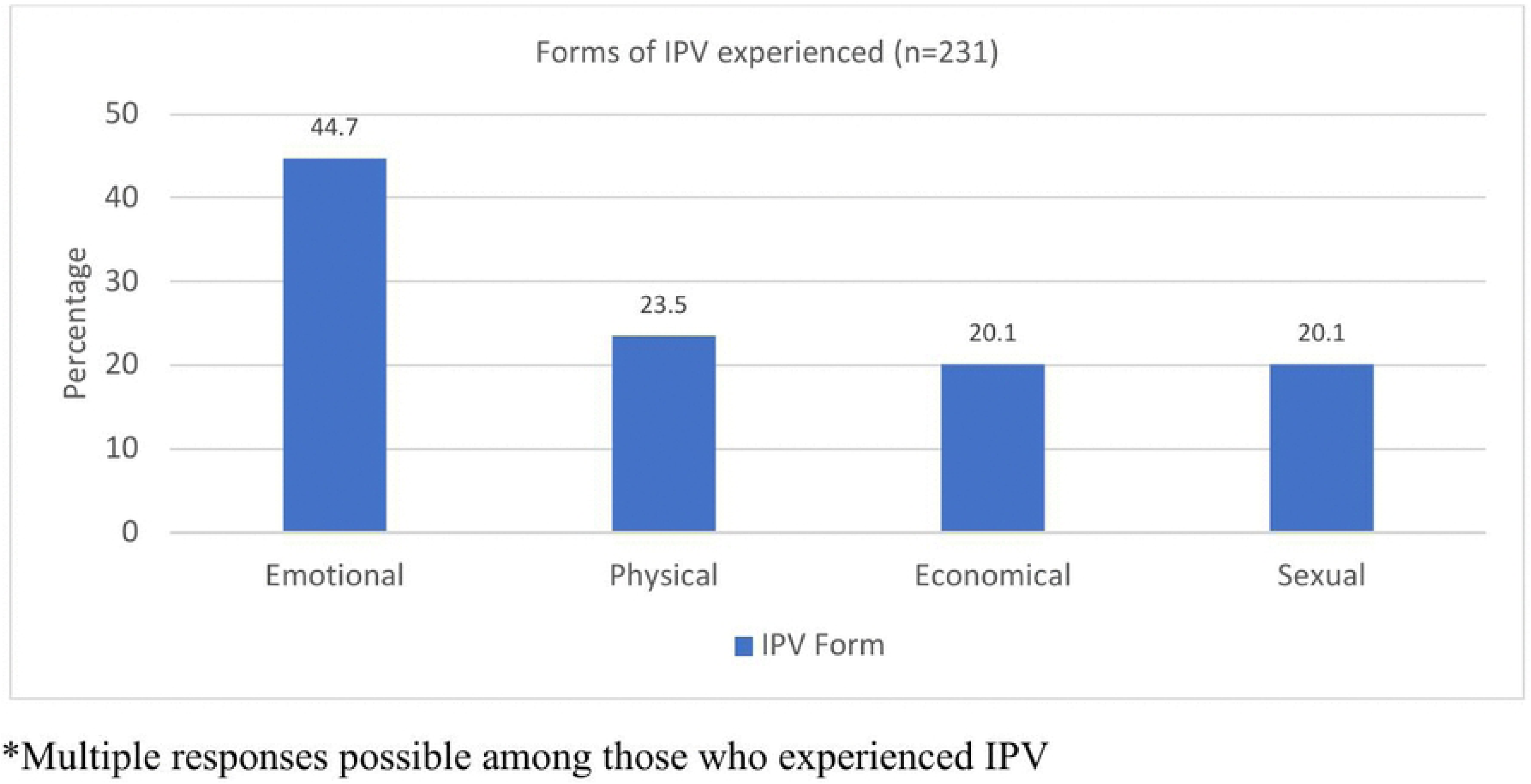
Forms of IPV experienced by women attending abortion care services at four selected hospitals in Lusaka district, Zambia, 2024 (n = 231)*

### Modern contraceptive use

Less than half (n=150, 41%) of the participants reported use of any modern contraceptive in the 6 months preceding the study, with half (n=75) of the users reporting use of injectables, 17% reporting use of OCPs (n=25), 13% reporting use of implants (n=19) and 19% (n=29) reporting use of male condoms. Although modern contraceptive use for all the methods studied was less among women who experienced IPV compared to those who did not experience it, the diferrence was seen more in the use of OCPs and male condoms.

**Fig 2.**
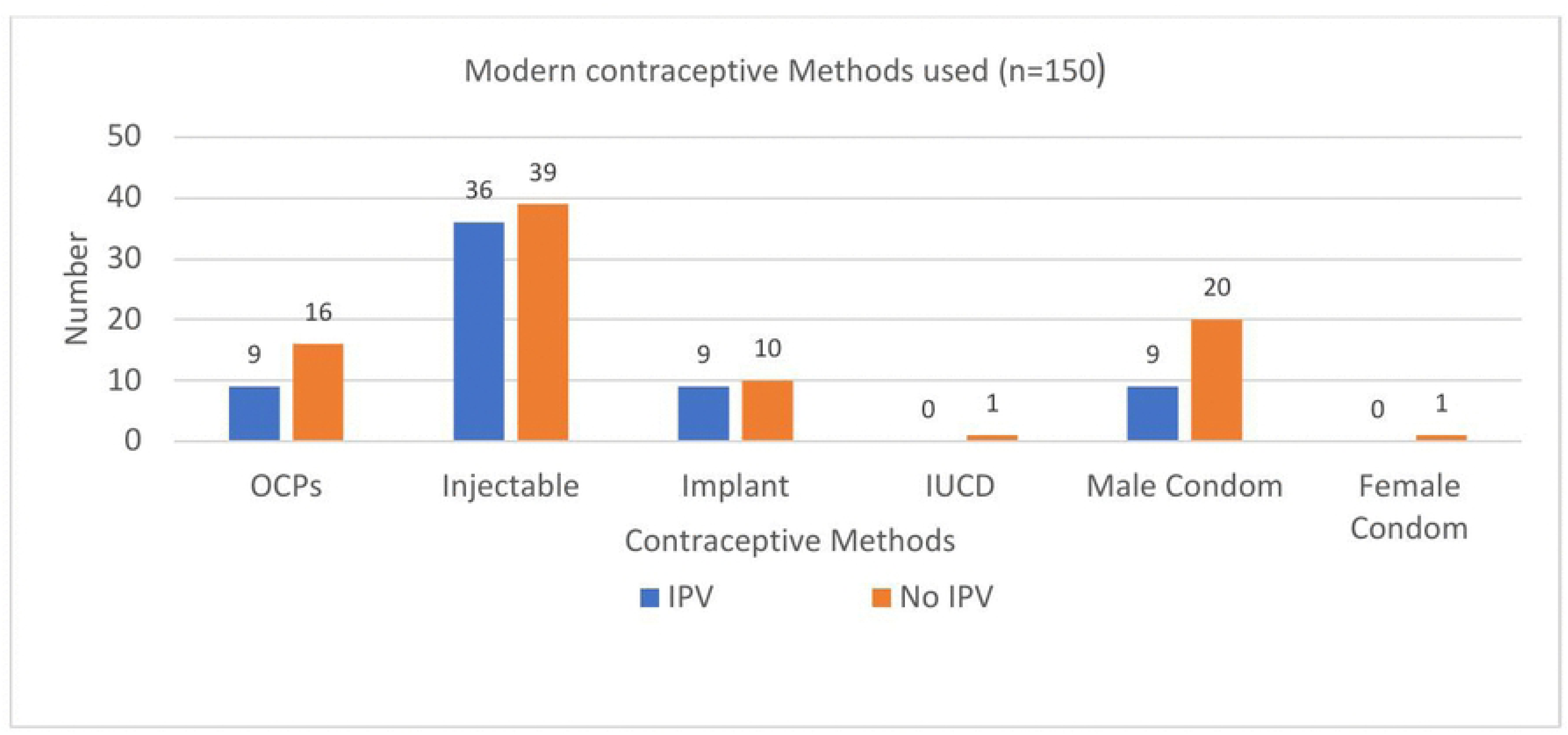
Proportions of modern contraceptive methods used by women who experienced IPV and those who did not experience IPV among those attending abortion care services at four selected hospitals in Lusaka district, Zambia, 2024 (n = 150)

### Factors associated with modern contraceptive use

On bivariate analysis, study site, experience of IPV, age category, marital status and parity were found to be significantly associated with modern contraceptive use (Table 1). In multivariable logistic regression, site, experience of IPV and age category remained significantly associated with modern contraceptive use (Table 2). Women who experienced IPV were less likely to use modern contraceptives compared to women who did not experience IPV (aOR 0.3; 95% CI, 0.17 – 0.52). The odds of modern contraceptive use among women seeking abortion care services from Chawama FLH were 4 times higher compared to women seeking abortion care services from the WNUTH (aOR 4.4; 95% CI 1.68 – 11.65). Women aged 25 year and above were 14 times more likely to use modern contraceptives compared to AGYW aged 14-24 years (aOR 14; 95% CI, 6.40 – 30.60).

**Table 2:** Multivariate logistic regression for factors associated with contraceptive use among women seeking abortion care services at four selected hospitals in Lusaka, Zambia, 2024 (n=367).

| Variable | <sup>a</sup>cOR | 95% <sup>b</sup>CI | p-value | <sup>c</sup>aOR | 95% CI | p-value |
| --- | --- | --- | --- | --- | --- | --- |
| **Age in Years** |  |  |  |  |  |  |
| 14-24 | <sup>d</sup>Ref |  |  |  |  |  |
| 25+ | **12.81** | **7.28 – 22.53** | **<0.0001** | **14.0** | **6.40 – 30.60** | **<0.0001** |
| **Site** |  |  |  |  |  |  |
| WNH-UTH | Ref |  |  |  |  |  |
| Chawama FLH | **3.20** | **1.37 – 7.47** | **0.007** | **4.42** | **1.68 – 11.65** | **0.003** |
| Kanyama FLH | **5.43** | **2.34 – 12.61** | **<0.0001** | **8.88** | **3.35 – 23.54** | **<0.0001** |
| Chipata FLH | **6.36** | **2.72 – 14.86** | **<0.0001** | **20.8** | **7.32 – 59.02** | **<0.0001** |
| **Experience of IPV** |  |  |  |  |  |  |
| No | Ref |  |  |  |  |  |
| Yes | **0.21** | **0.13 – 0.33** | **<0.0001** | **0.30** | **0.17 – 0.52** | **<0.0001** |
| **Marital status** |  |  |  |  |  |  |
| Married | Ref |  |  |  |  |  |
| Not married | **0.39** | **0.25 – 0.62** | **<0.0001** | 2.01 | 1.01 – 4.03 | 0.058 |
| **Parity** |  |  |  |  |  |  |
| Nullipara | Ref |  |  |  |  |  |
| Multipara | **4.0** | **2.51 – 6.32** | **<0.0001** | 1.84 | 0.90 – 3.77 | 0.096 |
| Grandmultipara | **6.7** | **1.60 – 28.1** | **0.009** | 2.02 | 0.39 – 10.57 | 0.403 |

<sup>a</sup>cOR = Crudes Odds Ratio, <sup>b</sup>CI = Confidence Interval, <sup>c</sup>aOR = Adjusted Odds Ratio, <sup>d</sup>Ref = Reference Category

## Discussion

The prevalence of IPV among women seeking abortion care services at the four hospitals in Lusaka district was 63% with emotional violence being the commonest form of IPV experienced. Modern contraceptive use among these women stood at 41%, with the commonest modern contraceptive method used being the injectable.

The 63% prevalence of IPV among women seeking abortion care services which was found in this study is higher than the 44% reported among women of reproductive age in East Africa [28] and the 47% reported among women of reproductive age in Zambia [8]. The difference in magnitude might be due to the fact that both studies involving women of reproductive age in East Africa and Zambia used findings from the demographic health surveys (DHSs). In the East African study, forms of IPV which were included were physical, sexual and emotional violence, while the Zambian DHS of 2018 included only physical and sexual forms of IPV. However, this study included four forms of IPV with the addition of economic IPV. In addition to the increase in the number of IPV forms in this study, the higher prevalence of IPV which was found may also be due to the fact that the study population was women seeking abortion care services who are at high risk of experiencing IPV [10]. A study in Sweden found that 29% of women seeking abortion services experienced IPV compared to 22% of women seeking contraceptive counselling [29]. The other explanation for the high prevalence of IPV among women seeking abortion care services which was found in this study could be that women who experience IPV may not be allowed to decide whether or not they use contraception and therefore choose to terminate an unwanted pregnancy.

With respect to the forms of IPV experienced, the prevalence for physical and sexual violence found in this study are consistent with findings from a study done in Sub-Sahara Africa (SSA) where the prevalence for physical violence was found to be 30% and that for sexual violence 13% [30]. However, the 45% prevalence of emotional violence found in this study was higher than the 30% which was found in SSA [30] and the 24% which was found in Ethiopia [31]. The high prevalence of emotional violence which was found in this study compared to what has been found in other studies may also be due to variations in screening questions used in different studies. For instance, in the study done by Turiye and colleagues in Ethiopia, the prevalence of emotional violence was found to 24% but in the same study, 57% of the women were said to have experienced “controlling behaviour” which has some overlap with emotional violence. The 20% prevalence of economic violence found in this study is comparable to the 21% which was found in Sudan but higher than the 4% found in Namibia and lower than the 51% found in Nigeria [32]. The wide range of prevalence rates in economic violence may be due to variations in definitions of economic violence and the various tools used to screen for economic violence across the globe. It has been noted that there is a lack of consistency about definition of economic violence and that there is no agreed upon index with which to measure it [6].

Modern contraceptive use among women in this study was found to be 41%. This figure is lower compared to the 48% modern contraceptive use among married women of reproductive age in Zambia [8]. It is however consistent with the 41% modern contraceptive use which was found among women seeking abortion services in Ethiopia [26]. The injectable was found to be the most common contraceptive method used by half of women were using modern contraceptives within six months prior to their participation in this study. This finding together with 17% of the users who were using OCPs are similar to the findings of the ZDHS of 2018 which found that 54% of the users were using the injectables and 17% were using the OCPs. However, this study found that the 13% of modern contraceptive users who were using the implant prior to conception was less than the 17% which was reported among married women in the ZDHS 2018 [8].

In this study, women who reported experiencing any form IPV were found to be less likely to use modern contraceptives compared to women who did not report experiencing any form of IPV. This finding is similar to the findings of previous studies in Africa and other parts of the world which have shown that the experience of IPV is associated with reduced modern contraceptive use among women in the general population [3, 33], as well as women seeking abortion services [34]. Various reasons may underlie the significantly low rate of modern contraceptive use among women who experience IPV. Women exposed to IPV usually lack control over their own sexuality. They may not be allowed whether or not they use modern contraceptives and so, they avoid using contraceptives all together for fear of violence. In contrast, a study involving Eastern Sub-Saharan (SSA) countries showed an opposite relationship between modern contraceptive use and IPV, with women who experienced IPV being more likely to use modern contraceptives [4]. One possible explanation advanced for the higher usage of modern contraceptives among women in the Eastern SSA study was that women in abusive relationships may attempt to prevent pregnancy because they do not want to bring a child into a violent family setting.

In our study, Women seeking abortion care services from Chawama FLH were found to be more likely to use modern contraceptives compared to women seeking services from the WNUTH. Women seeking services from Chawama FLH, which is located in a high density peri-urban area of Lusaka are likely to be less educated and unemployed compared to women seeking services from the WNUTH which is located in the urban area of Lusaka. However, in bivariate analysis, the level of education and employment status of women were found not to be significantly associated with modern contraceptive use. Women aged 25 years and above were more likely to use modern contraceptives compared to AGYW. This finding is consistent with finding from previous studies which have shown that modern contraceptive use is higher among older women compared to AGYW [20, 35]. The low levels of modern contraceptive use among AGYW may be due to inadequate knowledge among adolescents on the availability of sexual reproductive health services including contraceptive services, socio-cultural norms and mistrust of health workers [36].

In conclusion, women presenting for abortion care services at selected health facilities in Lusaka district experienced high levels of intimate partner violence and they had a low rate of modern contraceptive utilisation. Among the four forms of IPV experienced, emotional violence was the most common form. Women who experienced IPV, AGYW and women seeking services from the WNUTH were less likely to use modern contraceptives compared to those who did not experience IPV, older women and those seeking services from Chawama FLH respectively. Thus, policies and programmes aimed at increasing modern contraceptive use among women should pay particular attention at addressing IPV, including economic violence and the reproductive health needs of AGYW and women residing in peri-urban areas.

### Strengths and limitations

The strength of this study is that it included the economic form of IPV, in addition to the three forms which are usually reported in many studies on IPV. The study was conducted at four health facilities located in Lusaka district, which included the WNUTH located within the low-density residential areas, and three FLHs serving high density residential areas in Lusaka. Despite these strengths, it is worth acknowledging the limitations inherent in this study. Firstly, the study participants were women accessing abortion care services, and hence, generalisation to the wider population of women in the reproductive age has to be taken with caution. The use of multivariate regression in this study helped to reduce confounding to some extent. Secondly, both the experience of IPV and modern contraceptive use within six months prior to participation in the study were self-reported and therefore prone to recall and information bias. Also, there was a possibility of social desirability bias arising from the sensitive nature of questions on IPV and modern contraceptive use, which was reduced by holding interviews in an enviroment that ensured confidentiality and by using trained female interviewers.

## Data Availability

The data related to this study are available upon request. Due to ethical restrictions, the data underlying the study can not be made public. However, it is possible to obtain access to the dataset by submitting a reasonable request to the Directorate of Research and Development (DRD) at the University of Zambia (UNZA). The address for UNZA DRD is P.O Box 32379, Lusaka Zambia.

## Acknowledgements

Pre-Publication Support Services (PREPSS) supported the development of this manuscript by providing author training as well as pre-publication peer review.

## References

[1] United Nations. Sustainable Development Goals and Targets. 2020. Available from: https://zambia.un.org/sites/default/files/2020-10/sustainable_development_goals_and_targets_booklet.pdf

[2] World Health Organization. Family Planning/Contraceptive Methods. WHO Factsheet. 2025. Available from: https://www.who.int/news-room/fact-sheets/detail/family-planning-contraception

[3] Tomar S, Dehingia N, Dey AK, Chandurkar D, Raj A, Silverman JG. Associations of intimate partner violence and reproductive coercion with contraceptive use in Uttar Pradesh, India: How associations differ across contraceptive methods. PLOS ONE; 15. 16 October 2020. doi: 10.1371/journal.pone.0241008.

[4] Muluneh MD, Francis L, Agho K, Stulz V. The association of intimate partner violence and contraceptive use: a multi-country analysis of demographic and health surveys. Int J Equity Health; 22. 26 April 2023. doi: 10.1186/s12939-023-01884-9.

[5] White M, Fjellner D. The Prevalence of Economic Abuse Among Intimate Partners in Alberta. Sage Open; 12. January 2022. doi: 10.1177/21582440221084999.

[6] Postmus JL, Hoge GL, Breckenridge J, Sharp JN, Chung D. Economic Abuse as an Invisible Form of Domestic Violence: A Multicountry Review. Trauma Violence Abuse 2020; 21: 261–283.

[7] Sardinha L, Maheu-Giroux M, Stöckl H, Meyer SR, García-Moreno C. Global, regional, and national prevalence estimates of physical or sexual, or both, intimate partner violence against women in 2018. The Lancet 2022; 399: 803–813.

[8] Zambia Statistics Agency, Ministry of Health, ICF. Zambia Demographic and Health Survey 2018. DHS Final Reports FR361, Lusaka, Zambia, and Rockville, Maryland, USA: Zambia Statistics Agency, Ministry of Health, and ICF. Available from: https://dhsprogram.com/pubs/pdf/fr361/fr361.pdf

[9] Roth L, Sheeder J, Teal SB. Predictors of intimate partner violence in women seeking medication abortion. Contraception 2011; 84: 76–80.

[10] Hailu HT, Mekonnen W, Gufue ZH, Weldegebriel SG, Dessalegn B. Intimate partner violence as a determinant factor for spontaneous abortion during pregnancy: an unmatched case–control study. Front Public Health; 11. 5 June 2023. doi:10.3389/fpubh.2023.1114661.

[11] Kupoluyi JA. Intimate partner violence as a factor in contraceptive discontinuation among sexually active married women in Nigeria. BMC Womens Health 2020; 20: 128.

[12] Ahinkorah BO, Budu E, Aboagye RG, et al. Factors associated with modern contraceptive use among women with no fertility intention in sub-Saharan Africa: evidence from cross-sectional surveys of 29 countries. Contracept Reprod Med 2021; 6: 22.

[13] Baritwa MS, Joho AA. Intimate partner violence influences modern family planning use among married women in Tanzania: cross-sectional study. BMC Public Health; 9 February 2024. doi: 10.1186/s12889-024-17666-z.

[14] Drew LB, Mittal M, Thoma ME, Harper CC, Steinburg JR. Intimate Partner Violence and Effectiveness Level of Contraceptive Selection Post-Abortion. J Womens Health 2020; 29: 1226–1233.

[15] Fan X, Vignau Loria M. Intimate partner violence and contraceptive use in developing countries: How does the relationship depend on context? Demogr Res 2020; 42: 293– 342.

[16] Pallitto CC, García-Moreno C, Jansen HAFM, Heise L, Ellsberg M, Watts C. Intimate partner violence, abortion, and unintended pregnancy: Results from the WHO Multi-country Study on Women’s Health and Domestic Violence. Int J Gynecol Obstet 2013; 120: 3–9.

[17] Pearson E, Andersen KL, Biswas K, Chowdhury R, Sherman SG, Decker MR. Intimate partner violence and constraints to reproductive autonomy and reproductive health among women seeking abortion services in Bangladesh. Int J Gynecol Obstet 2017; 136: 290–297.

[18] Ahinkorah BO, Ameyaw EK, Seidu A-A. Socio-economic and demographic predictors of unmet need for contraception among young women in sub-Saharan Africa: evidence from cross-sectional surveys. Reprod Health 2020; 17: 163.

[19] Ahissou NCA, Benova L, Delvaux T, Gryseels C, Dossou JP, Goufodji S, et al. Modern contraceptive use among adolescent girls and young women in Benin: a mixed-methods study. BMJ Open 2022; 12: e054188.

[20] Achana FS, Bawah AA, Jackson EF, Welaga P, Awine T, Asuo-Mante E, et al. Spatial and socio-demographic determinants of contraceptive use in the Upper East region of Ghana. Reprod Health 2015; 12: 29.

[21] Islam A, Mondal N, Khatun L, Rahman M, Islam R, Mostofa G, et al. Prevalence and Determinants of Contraceptive use among Employed and Unemployed Women in Bangladesh. Int J MCH AIDS IJMA 2016; 5: 92–102.

[22] Makola L, Mlangeni L, Mabaso M, Chibi B, Sokhela Z, Silimfe Z, et al. Predictors of contraceptive use among adolescent girls and young women (AGYW) aged 15 to 24 years in South Africa: results from the 2012 national population-based household survey. BMC Womens Health; 19. December 2019. doi: 10.1186/s12905-019-0861-8.

[23] Zambia Statistics Agency. Zambia 2022 census of population and housing report. Lusaka, Zambia. Available from: https://www.zamstats.gov.zm/wp-content/uploads/2023/12/2022-Census-of-Population-and-Housing-Preliminary.pdf

[24] Festin MPR, Kiarie J, Solo J, Spieler J, Malarcher S, Van-Look PF, et al. Moving towards the goals of FP2020 — classifying contraceptives. Contraception 2016; 94: 289–294.

[25] Mayer M, Snow N, Haileyesus M, Tibebu D. Economic Abuse Screening Tool (EAST): A Toolkit for Social Service Providers. 2023. Available from: https://ccfwe.org/economic-abuse-screening-tool/

[26] Alemu L, Ambelie YA, Azage M. Contraceptive use and associated factors among women seeking induced abortion in Debre Marko’s town, Northwest Ethiopia: a cross-sectional study. Reprod Health 2020; 17: 97.

[27] Deshpande NA, Lewis-O’Connor A. Screening for Intimate Partner Violence During Pregnancy. Rev Obstet Gynecol 2013; 6: 141–147.

[28] Tessema ZT, Gebrie WM, Tesema GA, Alemneh TS, Teshale AB, Yeshaw Y, et al. Intimate partner violence and its associated factors among reproductive-age women in East Africa: A generalized mixed effect robust poisson regression model. PLOS ONE;18. 18 August 2023. doi: 10.1371/journal.pone.0288917.

[29] Öberg M, Stenson K, Skalkidou A, Heimer. Prevalence of intimate partner violence among women seeking termination of pregnancy compared to women seeking contraceptive counseling. Acta Obstet Gynecol Scand 2014; 93: 45–51.

[30] Mossie TB, Mekonnen Fenta H, Tadesse M, Tadele A. Mapping the disparities in intimate partner violence prevalence and determinants across Sub-Saharan Africa. Front Public Health 2023; 11: 1188718.

[31] Tiruye TY, Chojenta C, Harris ML, Holliday E, Loxton D. Intimate partner violence against women and its association with pregnancy loss in Ethiopia: evidence from a national survey. BMC Womens Health; 20. December 2020. doi: 10.1186/s12905-020-01028-z.

[32] Surviving Economic Abuse. Economic Abuse: A Global Perspective. 2022. Available from: https://survivingeconomicabuse.org/wp-content/uploads/2022/11/SEA_Economic-Abuse-A-Global-Perspective.pdf

[33] Zemlak JL, Marineau L, Willie TC, Addison H, Edwards G, Kershaw T, et al. Contraceptive Use Among Women Experiencing Intimate Partner Violence and Reproductive Coercion: The Moderating Role of PTSD and Depression. Violence Against Women 2024; 30: 2075–2095.

[34] Silverman J, Gupta J, Decker M, Kapur N, Raj A. Intimate partner violence and unwanted pregnancy, miscarriage, induced abortion, and stillbirth among a national sample of Bangladeshi women. BJOG Int J Obstet Gynaecol 2007; 114: 1246–1252.

[35] Demissie KA, Jejaw M, Teshale G, Tiruneh MG, Tafere TZ, Hagos A. Multinomial multilevel analysis of factors affecting the use of modern contraceptives in sub-Saharan Africa: evidence from 2015 to 2023 Demographic Health Survey. Sci Rep 2025; 15: 16751.

[36] Ministry of Health. Zambia National Adolescent Health Strategic Plan 2022-2026. 2022. Available from: https://www.unicef.org/zambia/media/5881/file/Zambia-National-Adolescent-Health-Strategic-Plan-2022-2026.pdf

